# Perceptions of Risk of Attending Hospital during the COVID-19 Pandemic: a UK public opinion survey

**DOI:** 10.1101/2020.08.24.20180836

**Authors:** Rebecca Pritchard, Emer M Brady, Yogini V Chudasama, Melanie J Davies, Gerry P McCann

## Abstract

In order to inform clinical and research practice in secondary care in light of the COVID-19 pandemic, an online survey was used to collect public opinions on attending hospitals. The survey link was circulated via the National Institute for Health Research (NIHR) Public Involvement (PPI) Leads network and social media. 402 people completed the survey. Participants age ranged from the 18-85+, with the majority (337 (84%)) aged between 35 to 74 years. There were a higher number of women (77%) compared to men (23%); and were mainly White European (91%) compared to BAME (6%), or other (2%).

Data collection included self-identified risk status due to comorbidity or age, and 100 point Likert-type scales to measures feelings of safety, factors affecting feelings of safety, intention to participate in research, comfort with new ways of working and attitudes to research.

Results for feelings of safety scales indicate two distinct groups; one of respondents who felt quite safe and one of those who did not. Implementation of COVID-19 related safety measures such as social distancing, use of PPE and cleaning were strongly supported by most respondents. There was ambivalence around less certain measures such as regular staff antigen and antibody testing. Respondents were most likely to participate in research related to their own condition, COVID-19 research and vaccine research, but less likely to participate in healthy volunteer research, especially if suffering from a pre-existing comorbidity identified with increased risk or were female. There was general agreement that participants are comfortable with new ways of working, such as remote consultation, though women and BAME respondents were less comfortable. Findings raise concerns for health inequalities already impacting some groups in the pandemic. The role of clinical necessity and personal benefit support the reopening of services in line with clinical necessity. Moderate caution in respect of vaccine research relative to patient-participant research presents a challenge for pending recruitment demands, and would benefit from qualitative research to explore themes and concerns in more depth and support development and targeting of key messaging.

**SUMMARY BOX:**

What is already known on this topic?
1. Very little is known about public perceptions of risk of exposure to COVID-19 and engagement with clinical and research provision in secondary care.
2. This research explores public perspectives in five key areas in order to inform health policy and both population and individual communication regarding attending secondary care sites for clinical and research activities.

What this study adds?
1. Insight into public risk perceptions specific to attending hospital during the COVID-19 pandemic
  a. There are distinct groups of people who do feel safe and those who do not
  b. Use of personal protective equipment, social distancing measures and cleaning are essential to supporting feelings of safety and are well supported
  c. Recruitment to vaccine and COVID-19 studies presents challenges, especially amongst women and BAME respondents
  d. Most people are very comfortable with new ways of working (i.e. remote/digital)
  e. There is very strong support for continued health science research
2. Insight into the differences in perceptions and attitudes by individual risk status (due to age or comorbidity), sex and ethnicity.

## INTRODUCTION

The novel infectious disease COVID-19, first identified in December 2019, has swept across the globe reaching pandemic levels with 12,685,374 confirmed cases and over half a million deaths.(1) It has led to significant changes in healthcare provision and clinical research activities associated with change in demand, practice and policy. In March 2020, the National Institute of Health Research (NIHR) Clinical Research Network suspended any new or ongoing studies at NHS and social care sites that were not nationally prioritised COVID-19 studies. The rapid reconfiguration of services meant many dedicated research personnel moved to the healthcare frontline, and remaining research personnel refocused work on COVID-19. Similar changes occurred in healthcare, with non-urgent clinical activity suspended and new, remote ways of working introduced to protect both patients and healthcare personnel as the UK entered a lockdown in March 2020. The UK began its vigilant journey out of lockdown in May 2020 and as it enters late summer the temporary halt of the many clinical and research activities in secondary and tertiary care will be ending. The adoption of new ways of working amid risks of initiating a second peak within the UK prompts a need for information on public opinion around attending hospital.

The importance of public involvement in all aspects of clinical and research activity is well recognised and enshrined in policy and procedures throughout health and social care; it’s relevance to the COVID-19 pandemic is reaffirmed by the NIHR (2). Public involvement improves the quality and relevance of research, (3) and though rapid escalation of research considering COVID-19 makes involvement more challenging these benefits are worth retaining. Public involvement can help researchers understand public perception of risk as the driver of a range of pandemic-related behaviours, (4) such as compliance with lockdown requirements, adoption of protective measures like mask wearing and social distancing, and wider engagement with services including health, screening, social care and education and therefore support communication efforts (5).

Understanding of public feelings of safety, perception of factors affecting feelings of safety, intention to participate in research, comfort with new ways of working and attitudes to research will support efforts to ease lockdown in the sensitive hospital environment and is a key component of NIHR Restart project (6). Procedures, and particularly communication, around restarting clinical and research activities within hospitals need to incorporate the public voice to maximise chances of success. Failure to ensure patients and research participants feel safe within the hospital could have wide ranging impact from failure to access necessary healthcare highlighted by the 57% decrease in A&E attendance in April 2020 (7), postponement or failure to access necessary treatment (8), successfully delivering key metrics for research outside of COVID-19, and failure to recruit to research essential to tackling COVID-19. Furthermore, communication and perception of risk is poorly understood and has potential to impact public trust in science (9).

We aimed to rapidly assess public attitudes to attending hospital across the UK for research purposes and clinical appointments.

## METHODS

### Study Participants

We developed an English language online survey in partnership with the NIHR Leicester Biomedical Research Centre Bioinformatics Hub in Research Electronic Data Capture Software (REDcap). The survey featured 1) participant information, 2) screening against inclusion/exclusion criteria and 3) survey questions. The survey was administered between 11 and 24 June, 2020, where lockdown restrictions were still in place across UK (England, Scotland, Wales and Northern Ireland). The link was shared via email and posted on social media (including websites, Twitter, and Facebook) by the NIHR Leicester BRC, the Leicester Diabetes Centre, the Centre for Black and Minority Ethnicity (BME) Health and NIHR PPI Leads nationally. Social media posts were sharable to facilitate snowball sampling.

### Study participants

The eligibility criteria were broad to maximise reach. We used the following inclusion criteria: 1) age 18 years or over; 2) resident in the UK; and 3) willingness to participate. Screening questions prevented completion in case of ineligibility

### Survey questions

Participant characteristics age, sex and ethnicity were collected, and whether they considered themselves classified at risk of COVID-19 because of a health condition (yes *vs*. no) or their age (yes *vs*. no). The questionnaire focused on perception of risk when attending hospitals during the pandemic. A 100-point Likert scale ranging from disagree (0) to agree (100) for each statement created by the researchers was presented with a simple interactive sliding scale. In total there were 42 statements: 11 explored current feelings of safety; 13 explored factors affecting feelings of safety; 4 explored intention to participate in research; 8 explored attitudes towards research; and 6 explored comfort with new ways of working. In addition, the opportunity to provide free text responses related to participants’ safety concerns was provided. The data from the free text fields are not included in this manuscript. Further details are provided in **Supplementary Methods S1**.

### Patient and public involvement

The survey was considered public involvement to inform the Leicester strategy for recommencement of clinical and research activities. It was soft launched to the Leicester PPI Groups, to gain initial data and feedback on any issues, with a full launch 3 days later. As no changes were required, the initial data in included in the full results.

### Ethics approval and informed consent

The survey included participant information, which remained accessible throughout survey completion. The screening questions included Boolean consent, as per best practice for remote consenting to non-interventional research. A clear explanation of the purpose of the survey, data handling, potential burden and benefits of participation was provided, and participants were prompted to carefully consider their willingness to participate.

This research has been reviewed by the Medicine and Biological Sciences Research Ethics Committee of the University of Leicester (ref:26258-rp237-ls:healthsciences).

### Statistical analysis

Descriptive analyses were performed. The continuous 100-point Likert scale was assessed for normality using histograms and the Shapiro-Wilk test. The distribution of all responses was found to be not normally distributed, therefore the median value and interquartile range (IQR) were used to present the findings. The responses were also stratified into four key groups: 1) whether or not the participant was classified at risk of COVID-19 because of a health condition; 2) or due to their age; 3) by men and women; and 4) White European or Black, Asian or Minority Ethnicity (BAME). The two-sample Wilcoxon rank-sum (Mann-Whitney) test was used to calculate whether there was a significant difference between groups, the statistical significance was set at P-value<0.05 (two-sided). Results are reported as median Likert scale followed by the interquartile range (median (IQR)). Participants were not required to answer every statement; thus, the total number of responses slightly vary due to missing data. Data from REDCap was exported into Stata version 16.0 to conduct data analyses.

## RESULTS

### Participants’ characteristics

A total of 402 participants completed the survey questionnaire. Of those, 192 (48%) reported they were at risk of the COVID-19 due to a health condition, and 286 (71%) due to their age. Participants age ranged from the 18-24 age group to 85+, where the majority (337 (84%)) were aged between 35 to 74 years. There were a higher number of women (77%) compared to men (23%); and were mainly White (91%) compared to BAME (6%), or other (2%), as shown in **Table 1**.

**Table 1.** Characteristics of participants who responded to the survey between 11 and 24 June, 2020 (N=402)

| Characteristics | No. (%) |
| --- | --- |
| **Risk of COVID-19 due to a health condition** |  |
| Yes | 192 (47.8) |
| No | 207 (51.2) |
| Missing | 3 (0.8) |
| **Risk of COVID-19 due to age** |  |
| Yes | 286 (71.1) |
| No | 109 (27.1) |
| Missing | 7 (1.7) |
| **Age group** |  |
| 18-24 | 7 (1.7) |
| 25-34 | 25 (6.2) |
| 35-44 | 52 (12.9) |
| 45-54 | 72 (17.9) |
| 55-64 | 98 (24.4) |
| 65-74 | 115 (28.6) |
| 75-84 | 30 (7.5) |
| 85+ | 3 (0.8) |
| **Sex** |  |
| Women | 308 (76.6) |
| Men | 94 (23.4) |
| **Ethnicity** |  |
| White | 366 (91.0) |
| BAME (Black and Minority Ethnicity) | 22 (5.5) |
| Other | 8 (2.0) |
| Missing | 6 (1.5) |

### Perception of attending hospitals

#### Current feelings of safety (11 statements)

Data are shown in Figure 1. Participants agreed they felt most safe and confident about coming to the hospital for essential surgery (median 78 (IQR 39-96)), followed by a clinical scan or x-ray, (median 77 (IQR 34-94)); and a clinical blood test (median 77 (IQR 35-94)). Whilst participants felt least safe and confident attending the Accident and Emergency (A&E) (median 50, IQR 21-85); or visiting a friend or family member in hospital, (median 49 (IQR 15-75)).

**Figure 1.**
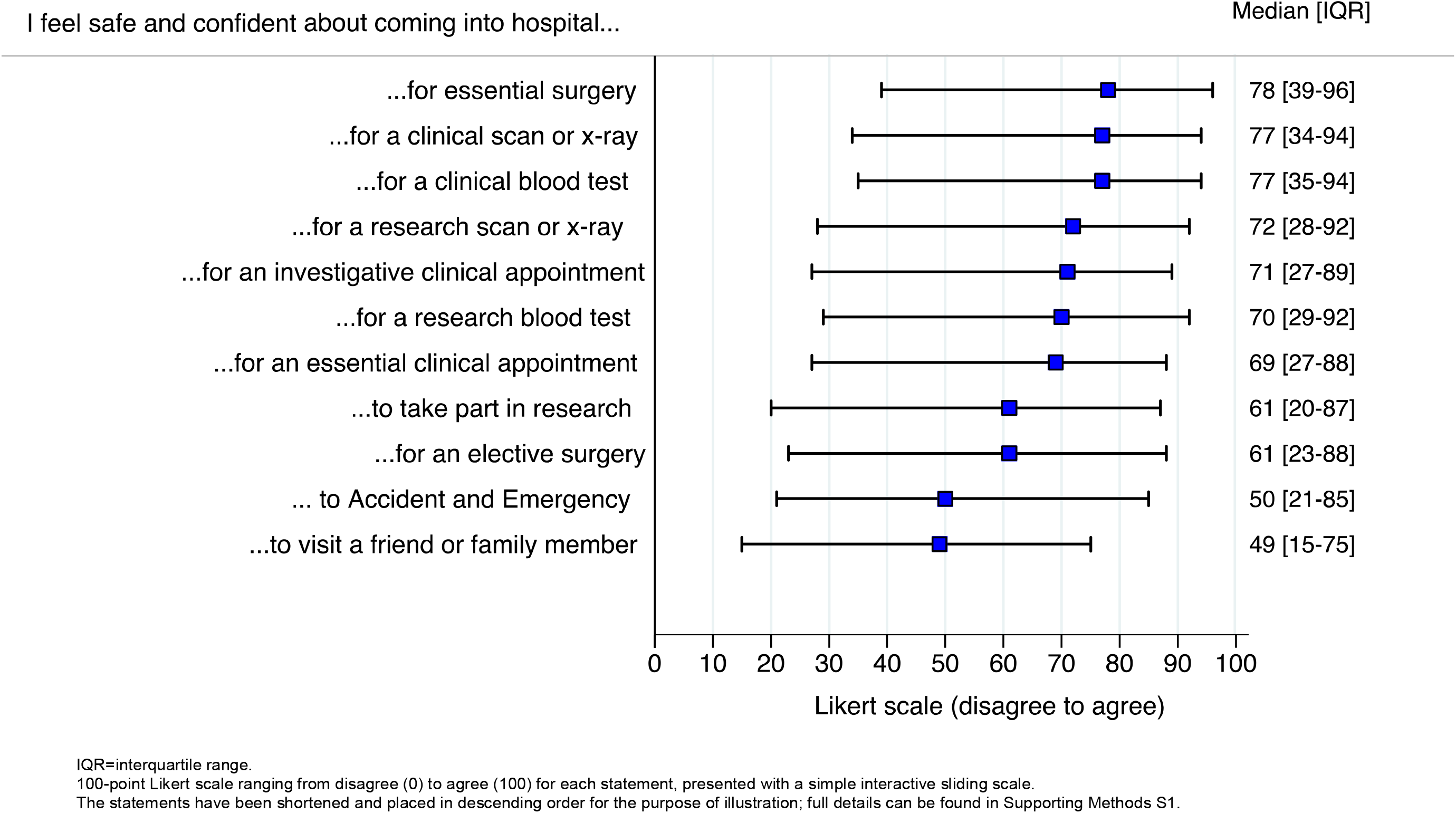
Participants response to their current feelings of safety when attending hospitals between 11 and 24 June, 2020

These findings significantly differed by those with a health condition compared to those without a health condition, as the responses on the Likert scale were much lower for all 11 statements. Particularly visiting a friend or family member in hospital with a health condition compared to those without a health condition, (median 33 (IQR 8-65) vs. 68 (IQR 32-90), respectively p<0.001), attending the A&E (median 36 (IQR 12-78) vs. 68 (IQR 33-90), P<0.001, respectively) or taking part in research (median 47 (IQR 12-81) vs. 73 (IQR 36-90), P<0.001, respectively). **Supplementary Table S1**. Similarly, women felt least safe and confident coming to the hospital compared to men, and those from a BAME background compared to those from a White European background. The BAME sample rated the Likert scale the lowest, even for coming to hospital for an essential clinical appointment (median 29 (IQR 21-57), P=0.003), investigative clinical appointment (median 35 (IQR 21-67), P=0.008), or clinical blood test (median 39 (IQR 18-70), P=0.028). There were no significant differences by participants who thought their age put them at increased risk of COVID-19 (**Supplementary Table S1**).

#### Factors affecting feelings of safety (13 statements)

In order to feel safe in the hospital environment the highest score on the Likert scale was to see consistent use of personal and protective equipment (PPE) such as gloves and masks, (median 95 (IQR 83-99)); to be reassured that careful cleaning measures were in place (median 95, (IQR 80-99)); to see strict social distancing measures in place (median 90 (IQR 76-98)); and to see as few staff as possible i.e. rather than seeing a doctor and having a nurse take a blood sample, everything is done by one person (median 82 (IQR 64-96)), as demonstrated in **Figure 2**. When stratifying the results, these statements were found to be mostly higher in those with a risk of the COVID-19 due to their health condition or age. Compared to the White European ethnicity, the BAME sample showed they felt safe in the hospital if they were reassured that staff have been tested negative for the COVID-19 infection (median 90 (IQR 77-95) vs. 76 (IQR 50-96), P=0.041, respectively), and also to attend research visits somewhere other than a hospital (median 89 (IQR 70-100) vs. 69 (IQR 49-87), P=0.006, respectively), **Supplementary Table S2**.

**Figure 2.**
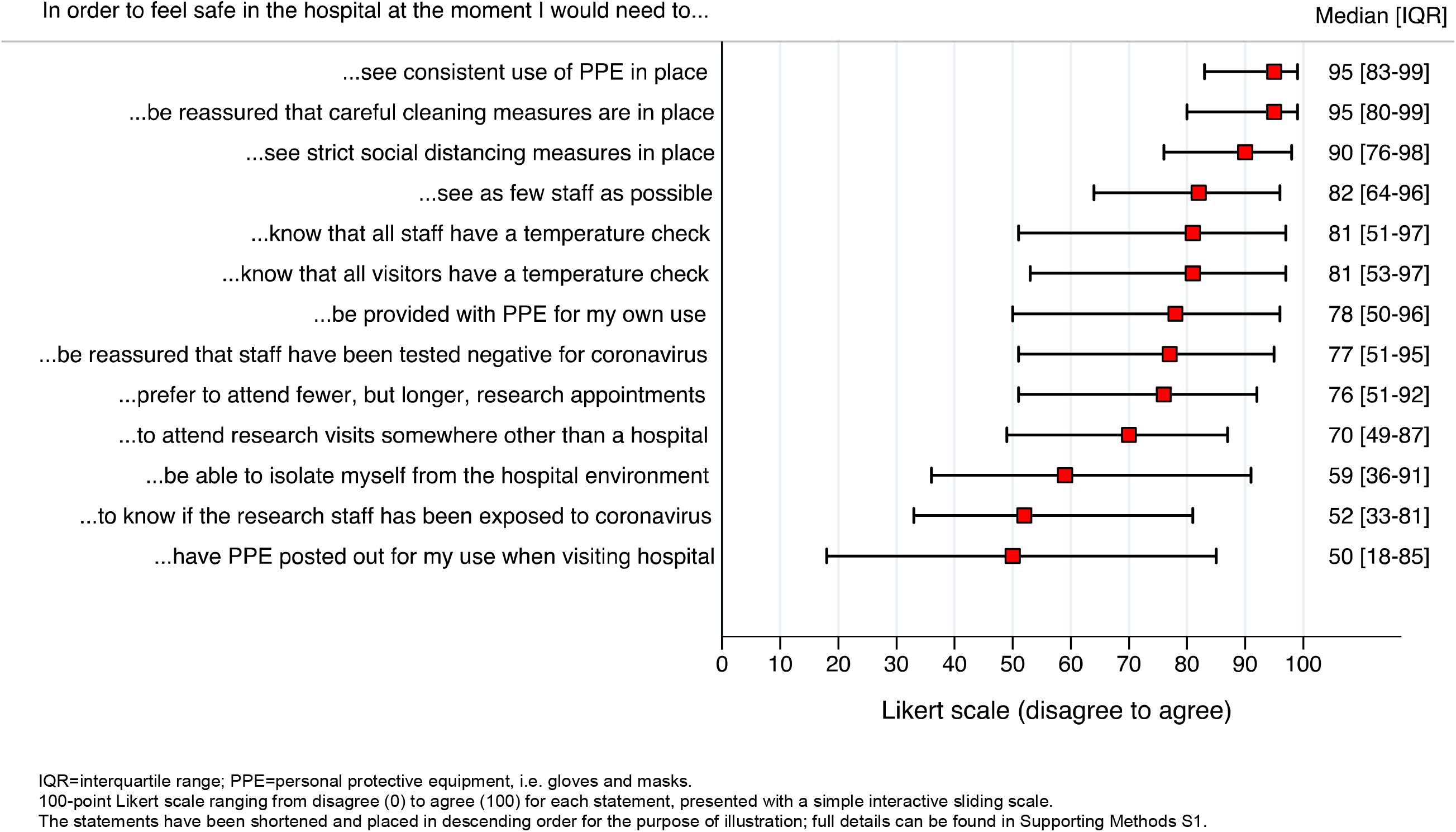
Participants response to factors affecting their feelings of safety when attending hospitals between 11 and 24 June, 2020

#### Intention to participate in research (4 statements)

Most participants would come into hospital to take part in research related to a medical condition (median 75 (IQR 49-96)); and in COVID-19 research which is not a vaccine study (median 70 (IQR 34-92)), Figure 3. The responses differed between men and women with men rating intention to participate in research more highly, and by risk of COVID-19 due to their health condition with those not at risk rating intention to participate in research more highly (**Supplementary Table S3**).

**Figure 3.**
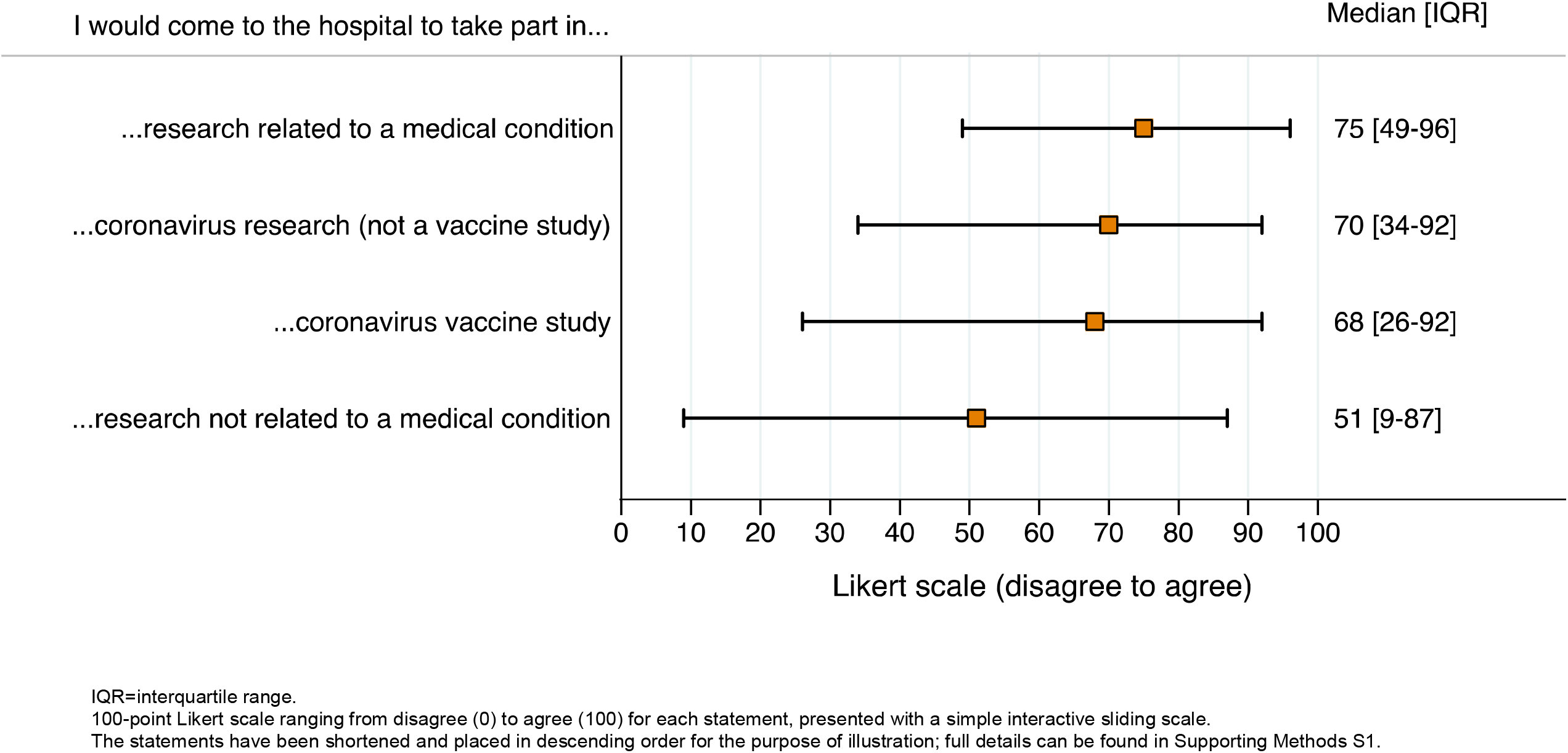
Participants response to their intention to participate in research between 11 and 24 June, 2020

#### Attitude towards research (8 statements)

Participants felt since the COVID-19 pandemic it is more important than ever to do health research (median 94 (IQR 79-99)), and also disagreed to the statements: we need to invest less money and resources in research (median 6 (IQR 2-21)); and were less interested in health science and research (median 6 (IQR 2-20)), as illustrated in **Figure 4**. Some of the statements varied by age, sex and ethnicity, **Supplementary Table S4**. This shows that those at risk due to age agreed that it is important to do research, disagreed with investing less in research, and disagreed they were less interested in health science and research more strongly than those not at risk due to age. Men disagreed more strongly with women on all statements about researchers asking participants into hospitals to do research. White participants agreed more strongly that it is important to do research, and disagreed more strongly that they are less interested in health and science, than BAME respondents.

**Figure 4.**
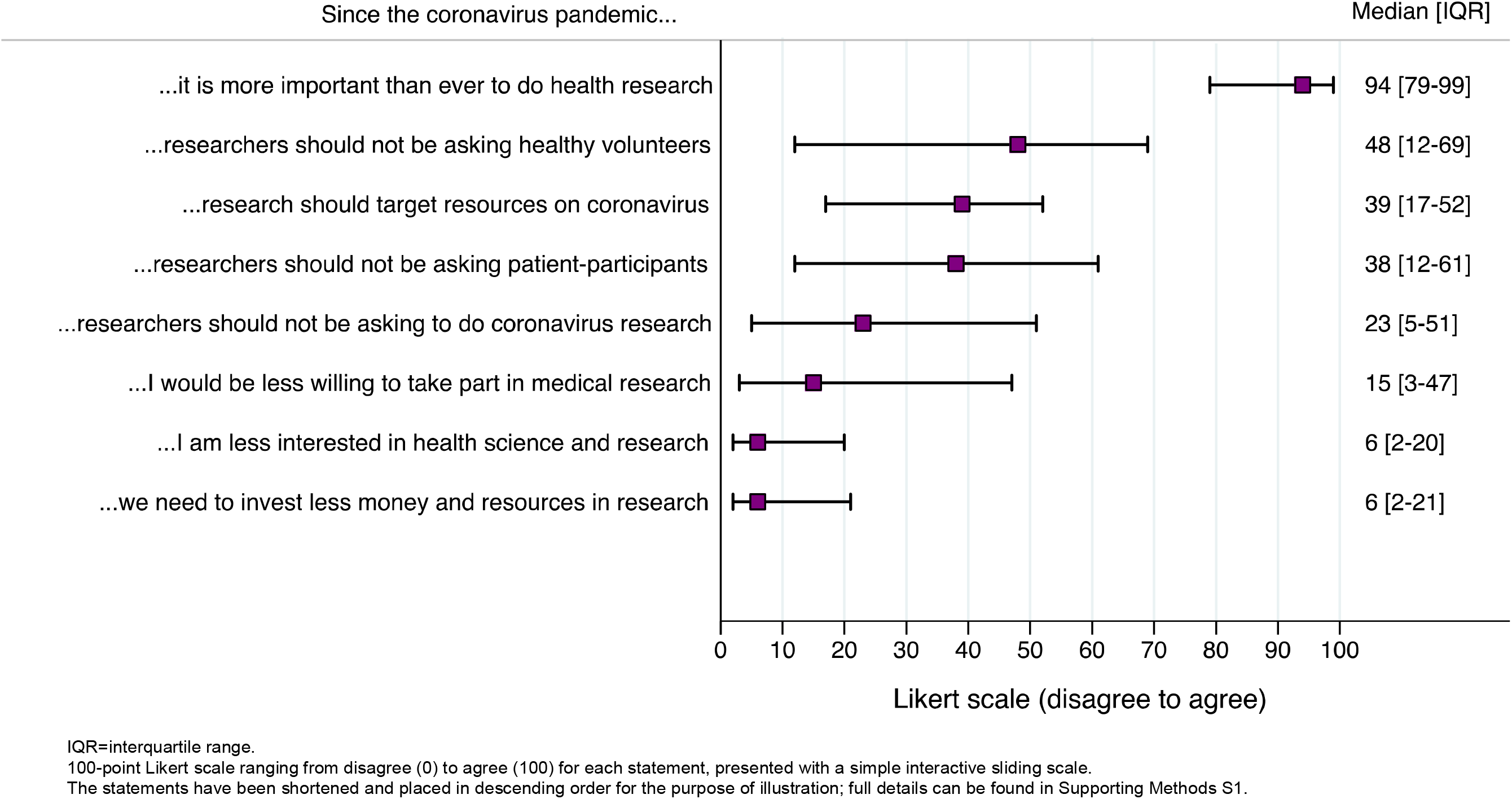
Participants response to their attitude towards research between 11 and 24 June, 2020

#### New ways of working (6 statements)

The results in **Figure 5** illustrates that participants were comfortable with new ways of working, as the results were very high for all statements. The 6 statements significantly differed between sex and ethnic groups (**Supplementary Table S5**). We found the responses were lower in BAME than those from a White background in the following statements: sharing my medical information, for research purposes using an online form or app (median 51 (IQR 11-90) vs. 95 (IQR 75-99), P<0.001), giving consent to take part in a research project using an online form or app (median 52 (IQR 24-89) vs. 96 (IQR 76-99), P<0.001, respectively), and women rated their comfort as lower compared to men across all scales, **Supplementary Table S5**.

**Figure 5.**
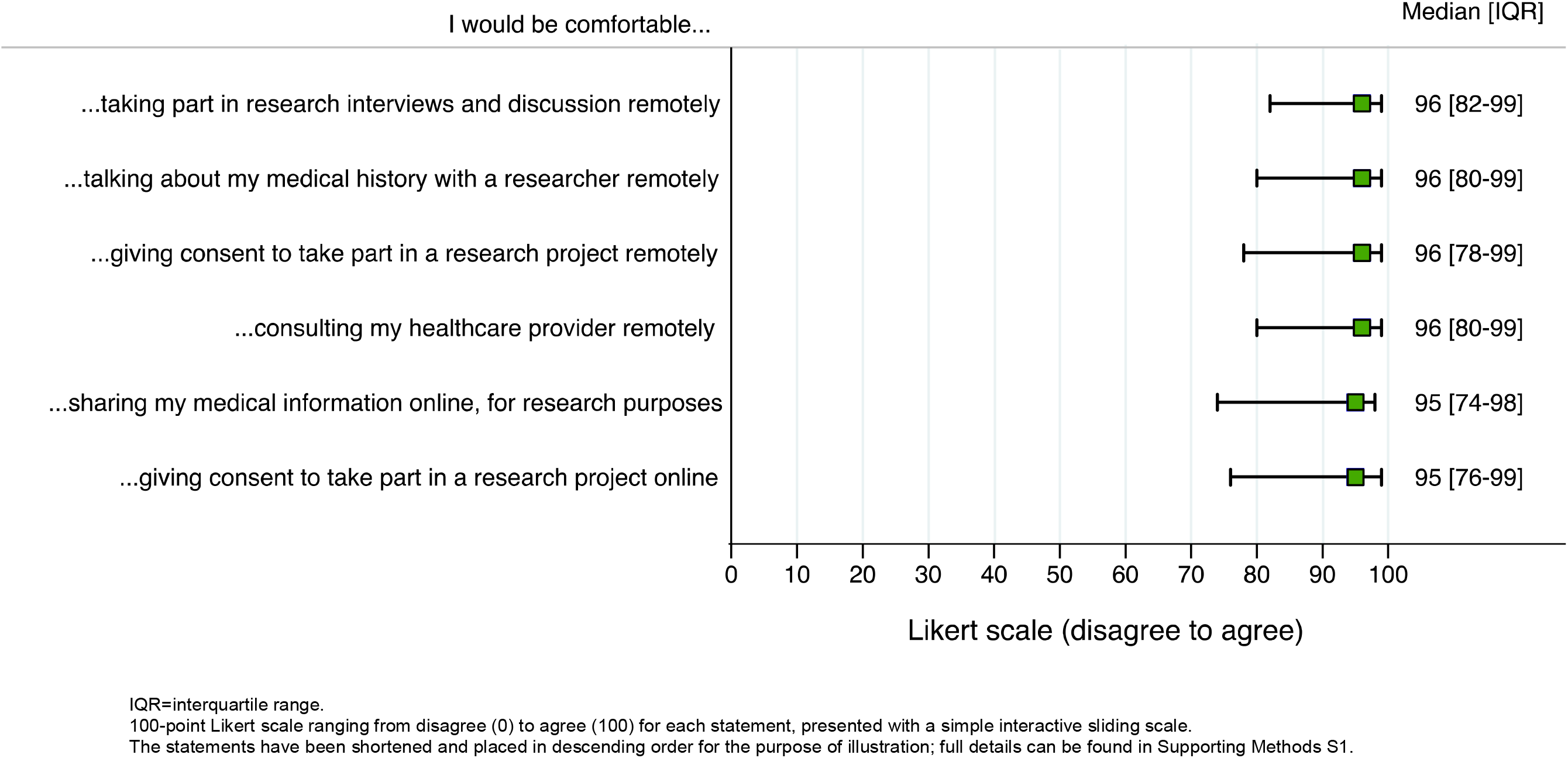
Participants response towards new ways of working between 11 and 24 June, 2020

## DISCUSSION

The purpose of this work was to rapidly assess public attitudes to attending hospital across the UK for research purposes and clinical appointments. The findings showed patterns of response that may support efforts to recommence clinical and research activity in secondary care. Of particular note are findings around differences between the perceptions and attitudes of women and BAME respondents suggesting a need to consider how current changes in activity might disproportionately impact some groups in society.

There is very little previous research into the risk perceptions associated with COVID-19, though our findings do reflect the paradoxical finding of (10) that men are more at risk than women (11), but women perceive greater risk than men. This is of particular interest as men are at greater risk but consistent with a large body of research showing women perceive greater risk than men across a range of activities (12). However, the lack of gender differences in respect of factors effecting feelings of safety would suggest the effect is underpinned by a requirement to see safety measures implemented i.e. the effect is a mix of the cognitive and emotive.

The differences between ethnic groups with lower scores for both feelings of safety and intention to participate in research and/or attend clinical appointments in our BAME responders is particularly relevant. The disproportionate risk of contracting COVID-19 (13) and poorer outcomes in BAME groups compared to white counterparts (14) is a recognised public health issue. Public Health England has engaged with key stakeholders to start the process of understanding this health inequality and discuss strategies to reduce the direct and indirect impact of this pandemic and indeed any future pandemic. The results presented here add to the growing evidence for the need to work with local communities to reduce fear and rebuild the BAME communities trust in the health services. Strategies must be sought to increase attendance for routine appointments need to be considered including increasing accessibility by bringing care to our BAME communities. Further, as new recruitment efforts for COVID-19 research commences; a focus on working with the BAME communities is required to permit adequate ethnic representation in health research because insufficient diversity in recruitment has consistently underpinned and exacerbated health inequalities. The lower feelings of comfort with new ways of working also highlights a potential area for further exacerbation of health inequality in service provision indicating services need to be patient-centred and offer choice of mode of contact.

The high perception of risk in attending Accident and Emergency (A&E), is notable and in line with recent findings (5). These results mirror what has been observed nationally with dramatic reduction in attendance to A&E and emergency admissions, April saw a staggering 57% drop compared to the same month in 2019 (7). The question is whether this is a positive change in public behavior or has this added to the indirect impact of COVID-19 on health. In, both scenarios work is required with the public and health systems to either continue diverting ‘treatment seeking’ away from A&E where it is not necessary or breakdown this new fear in seeking emergency care.

Histograms for scales (not shown) showed there was a distinct grouping of respondents into those who felt safe and those who did not, with generally few people in the middle. This was partially accounted for by differences associated with sex and ethnicity though interestingly there was no effect of age-related risk. Other factors may be associated with the bipolarity of responses, such as worldview, political inclination and sense of individualism, as identified in other recent research (10).

The factors effecting feelings of safety provide information on what participants expect to see when attending hospital. The highest rating for consistent use of PPE reinforces the recent decision to enforce use of PPE in hospitals by both visitors and staff in all areas. Findings suggest hospital attendees and particularly those at risk because of age/comorbidity will also need to see strict cleaning procedures and social distancing. BAME respondents additionally rated off site research visits and staff antibody and swab testing as important to their feelings of safety. In order to ensure representative recruitment to research and particularly rapid research around COVID-19, it will be important to consider how needs differ for potential BAME research participants in order to avoid perpetuating health inequalities.

Women and those at risk due to comorbidity were less likely to participate in research suggesting potential participants consider personal relevance of the research and societal urgency when deciding if they will participate. This highlights a specific recruitment challenge when considering vaccine trials for COVID-19 that will need to recruit people with comorbidities. Escalation of vaccine research will require large-scale public facing recruitment that has not been attempted previously in the UK.

The respondents overall attitude to research indicate a strong continued support for participation, interest and investment in health science research though ambivalence about prioritising COVID-19 suggests this is partially generic. The pandemic, and increased health science coverage in the media, provides an opportunity to increase engagement across the board, with age-related disparities suggesting there is a need to engage younger/working age populations. Respondents again balanced risk with personal and social necessity, finding it most acceptable to be asked to attend a hospital for COVID-19 research and least so for healthy volunteer studies, with a near-significant lower acceptability for those with comorbidity.

Communication professionals should consider pre-recruitment engagement and messaging in order to 1) prime a new audience for recruitment into vaccine studies that have typically relied on staff and student recruitment and 2) prime under-represented audiences with good quality information on risk and risk management to support recruitment efforts.

Finally, respondents reported high levels of comfort with digital and remote ways of working which is reassuring for clinicians and researchers. However, our aging and BAME communities needs considering given the reported differences in preferences.

A limitation of this work is the much smaller number of BAME respondents (6%) compared to White (91%) resultant from the need to be responsive this survey was undertaken rapidly, in just 2 weeks and only in English. Due to the limited nature of the sample therefore it is important to be cautious generalising especially as we found significant differences by ethnicity.

## CONCLUSIONS

We believe this is the first study in the UK to assess public opinions of attending hospitals during the rapid rise of the COVID-19 outbreak, which is particularly relevant to national activity around recommencing clinical and research activity.

As some of the most interesting findings pertain to groups under-represented in the sample it follows that further research into the thoughts and feelings of BAME communities and women would be informative. The patterns of risk perception suggest there may be complex processes underpinning individual assessment of risk, widely recognised as subjective (5), which might be explored more with qualitative research methodology. Focus groups are underway to explore this, and vaccine study recruitment, in more depth so diverse perspective can support both clinical and research activity post-COVID-19.

Healthcare needs to be accessible to BAME communities and women so alterations to practice need to take into account the differences in preference and include a flexible approach to the delivery of care.

## Data Availability

Data is available on request to the authors and completion of relevant ethical approval

## Funding source

There was no specific funding for this project. Staffing was funded through infrastructure funding for the NIHR Leicester BRC.

## Author contributions

Ranjit Arnold proposed consulting patients around hospital attendance for imaging and tests. RP, GM and EB developed the study design and survey in detail, expanding to include wider research and clinical concerns and aligned to the Restart project. Sue Sterland created the REDcap survey in consultation with RP. RP engaged the NIHR PPI Leads, local NIHR colleagues and University Hospitals of Leicester Trust to disseminate the link nationally, and via her own local PPI mailing lists. YC undertook statistical analysis and associated presentation of data. RP, EM, YC, GM worked on the manuscript collaboratively.

## Acknowledgements

The support of the NIHR PPI Leads Network with dissemination of the link for data collection is acknowledged.

## Data access and responsibility

The data is accessible to RP, SS, YC, EB and GM. It is the responsibility of RP.

## Declarations of interests

This activity may contribute to the PhD studies of RP.

No other interests are declared.

## Notes

### Competing Interest Statement

The authors have declared no competing interest.

### Funding Statement

No specific funding was granted for this piece of research. Staff was available through NIHR funding of the NIHR Leicester BRC.

